# Cardiac Ultrasonic Tissue Characterization in Myocardial Infarction Based on Deep Transfer Learning and Radiomics Features

**DOI:** 10.1101/2024.03.29.24305067

**Authors:** Ankush D. Jamthikar, Quincy A Hathaway, Kameswari Maganti, Yasmin Hamirani, Sabahat Bokhari, Naveena Yanamala, Partho P. Sengupta

## Abstract

**Objective:** Acute myocardial infarction (MI) alters cardiomyocyte geometry and architecture, leading to changes in the acoustic properties of the myocardium. This study examines ultrasomics — a novel cardiac ultrasound-based radiomics technique to extract high-throughput pixel-level information from images—for identifying infarcted myocardium.

**Methodology:** A retrospective multicenter cohort of 380 participants was split into two groups: a model development cohort (n=296; 101 MI cases, 195 controls) and an external validation cohort (n=84; 40 MI cases, 44 controls). Handcrafted and transfer learning-derived deep ultrasomics features were extracted from 2-chamber and 4-chamber echocardiographic views and ML models were built to detect patients with MI and infarcted myocardium within individual views. Myocardial infarct localization via texture features was determined using Shapley additive explanations. All the ML models were trained using 10-fold cross-validation and assessed on an external test dataset, using the area under the curve (AUC).

**Results:** The ML model, leveraging segment-level handcrafted ultrasomics features identified MI with AUCs of 0.93 (95% CI: 0.97-0.97) and 0.83 (95% CI: 0.74-0.89) at the patient-level and view-level, respectively. A model combining handcrafted and deep ultrasomics provided incremental information over deep ultrasomics alone (AUC: 0.79, 95% CI: 0.71-0.85 vs. 0.75, 95% CI: 0.66-0.82). Using a view-level ultrasomic model we identified texture features that effectively discriminated between infarcted and non-infarcted segments (p<0.001) and facilitated parametric visualization of infarcted myocardium.

**Conclusion:** This pilot study highlights the potential of cardiac ultrasomics in distinguishing healthy and infarcted myocardium and opens new opportunities for advancing myocardial tissue characterization using echocardiography.

## Introduction

Acute myocardial infarction (MI), defined pathologically as myocardial cell death due to prolonged ischemia, is one of the leading causes of morbidity and mortality globally. The prevalence of the disease approaches 3 million people worldwide, with 605,000 new MIs diagnosed annually in the US [1–3]. Echocardiography is a rapid, noninvasive, portable, and inexpensive imaging modality, making it the preferred technique for the assessment of MI patients [4, 5]. Although echocardiography visualizes the effects of ischemia and MI on regional and global myocardial function, direct identification and quantification of infarcted tissue remains challenging and has not been extensively explored, especially when compared to cardiac magnetic resonance (CMR) imaging, which is considered the gold standard for infarct tissue characterization [6].

The recent developments in image analysis and novel informatics approaches have augmented methods that can extract information from medical images that quantify their phenotypic characteristics in an automated, high-throughput manner. The application of such texture-based image analyses is referred to as ‘radiomics.’ Such features have been used recently to help prognosticate, predict treatment outcomes, and assess tissue malignancy in cancer research [7–9]. In neuroscience, radiomics have enabled the detection of Alzheimer’s disease [10, 11] and the diagnosis of autism spectrum disorder [30]. Although the ultrasound texture of the myocardium carries unique and specific information about the infarcted myocardium, cardiac ultrasound is not currently utilized clinically for myocardial tissue characterization; previous studies have reported that the intensity of the ultrasound backscatter is related to the physical properties of the myocardium [12, 13]. Moreover, there is limited information regarding the specific application of current informatics methods to enhance ‘radiomics-derived’ texture-based analyses for cardiac ultrasound imaging. Such features capture subtle, underlying information about the texture, shape, and intensity of structures within the heart, which may not be evident to the human eye, and therefore, enhance physicians’ comprehension by providing a deeper understanding of the underlying disease [14]. Our group has recently proposed the use of cardiac ultrasound radiomics, also referred to as “ultrasomics”, for accurately predicting cardiac remodeling and diastolic dysfunction using echocardiography imaging [15, 16].

In recent years, deep convolutional neural networks (CNNs) have been used extensively for segmentation, quantification, and diagnostic determinations using echocardiography images [17–19]. However, the successful application of deep learning requires large training samples. For smaller data sets, an approach using pre-trained CNNs known as “transfer learning” can be employed to avoid overfitting and replace deep learning in many practical applications. In this work, we compare both handcrafted and transfer learning-derived deep cardiac ultrasomics for the recognition of infarcted myocardium. We hypothesize that infarcted myocardium alters the myocardial texture and can be identified using cardiac ultrasomics in echocardiography frames, independent of regional and global LV function assessments. Figure 1 shows the central illustration of this study and also provides an overview of the MI prediction at the individual patient and the individual echocardiographic view.

**Figure 1.**
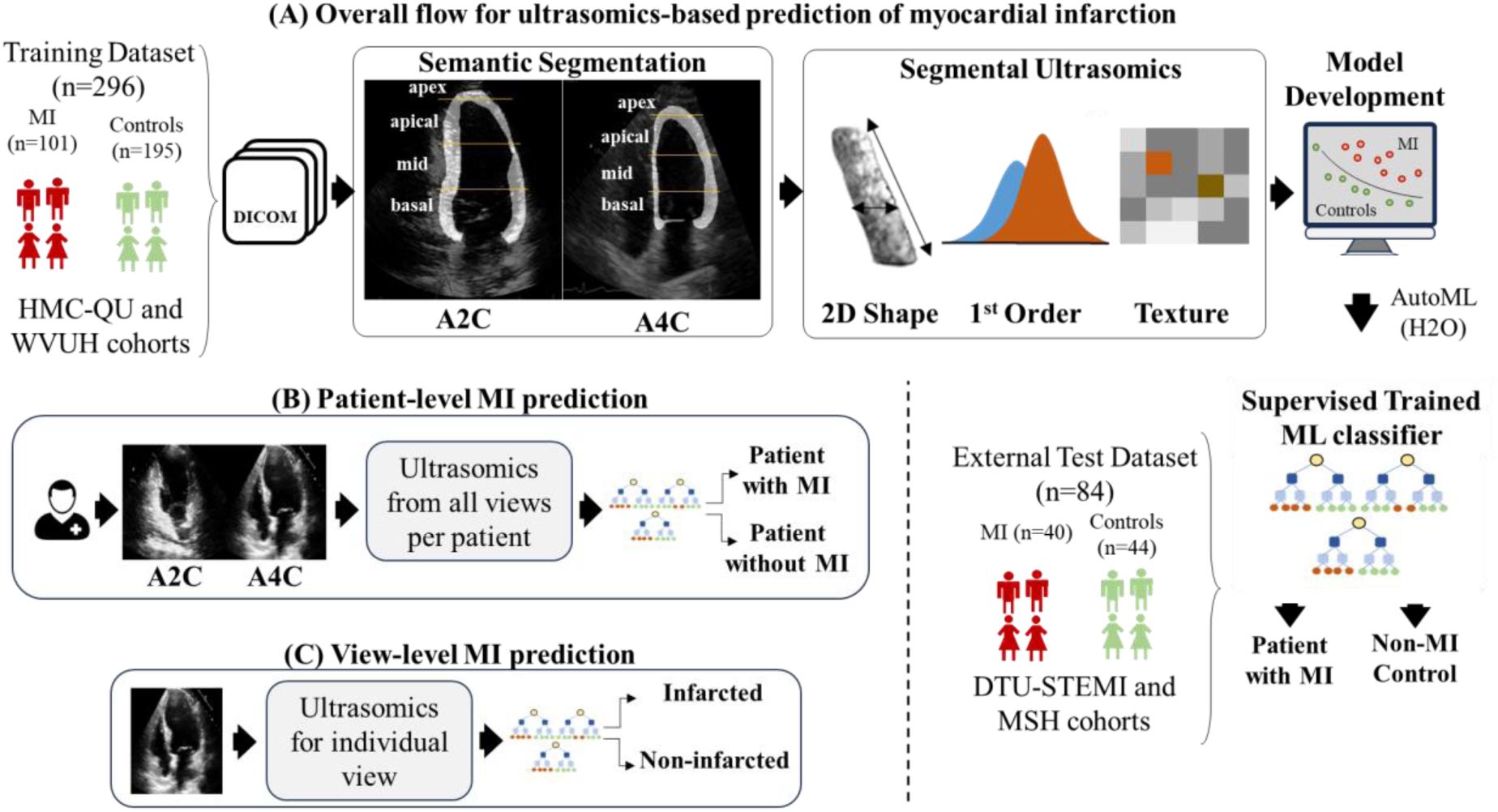
(A) Study design and central illustration indicating identification of MI and controls from the ultrasomics features extracted from 2-chamber and 4-chamber echocardiographic views, (B) predicting MI for an individual patient using a patient-level model, and (C) predicting infarcted echocardiographic view using a view-level model. HMC-QU = Hamad Medical College and Qatar University; WVUH = West Virginia University Hospital; DTU-STEMI = Door-To-Unload in ST-segment–elevation myocardial infarction; MSH = Mount Sinai Hospital; ML = machine learning; MI = myocardial infarction; A2C = 2-chamber view; A4C = 4-chamber view.

## Methodology

### Clinical population and study design

In this retrospective multicenter study, we used both open-source and core-lab databases with a total of 380 subjects, spanning four centers to develop diverse training and external validation cohorts that included both MI and control patients (Figure 1). The training cohorts consisted of 162 individuals (101 with MI and 61 non-MI controls) drawn from a publicly available database from Hamad Medical College and Qatar University (HMC-QU) [20, 21] and 134 non-MI controls recruited previously at the West Virginia University (WVU) without (n=57; healthy controls) and with (n=77) cardiovascular risk factors such as diabetes and hypertension [22]. The external validation cohort included 40 participants from the DTU-STEMI (Door-To-Unload in STEMI) trial—a prospective, multicenter, randomized pilot trial involving 14 centers in the United States [23] and 44 non-MI controls with risk factors and a normal CT coronary artery angiography recruited at Icahn School of Medicine at Mount Sinai [24]. These four cohorts have been described previously [21–24]. All four databases were approved by the respective local IRBs [21, 23, 25] and study participants provided written informed consent. The database collected from WVU, Mount Sinai, and DTU-STEMI studies was in accordance with the ethical standards of the institutional and national research committee and with the 1964 Helsinki Declaration.

### Defining the status of myocardial infarction

MI and other clinical variables were recorded in the HMC-QU echocardiography database [20, 21]. In addition, each echocardiographic video is provided with view-specific segment-level labels (17 myocardial AHA-defined segments [26]) that indicate the presence of disease and non-diseased segments. In the DTU-STEMI trial [23], patients underwent CMR imaging on days 3 to 5 and again on day 30 (±7 days). Standard protocols were used as detailed previously [23] with a central core laboratory (Duke Cardiovascular Magnetic Resonance Center, Durham, NC) performing CMR assessments on deidentified images. The presence of an infarct in each of the 17 myocardial AHA-defined echocardiography segments was labeled based on the presence of cardiac magnetic resonance-defined delayed hyperenhancement or late gadolinium enhancement (LGE) in each segment. In addition, we also considered delayed enhancement scores determined using the well-established 5-point grading system outlined by the AHA [27], where 0 indicates no hyperenhancement, 1 represents 1%–25%, 2 signifies 26%–50%, 3 corresponds to 51%–75%, and 4 reflects 76%–100% involvement [27].

### Echocardiography image analysis and segmentation

Our automated echocardiography imaging workflow is comprised of four stages: preprocessing, view identification, segmentation, and ultrasomics feature extraction. In the preprocessing stage, to ensure uniformity and compatibility, 2D echocardiograms from various formats (.avi and mp4) were converted into the DICOM format using the proprietary Sante DICOM software. DICOM files containing Doppler data or dual ultrasound regions were excluded. The view identification stage used these processed multi-beat echocardiogram DICOM files to classify the transthoracic echocardiography views allowing identification of apical 4-chamber (a4c) and apical 2-chamber (a2c). During the segmentation stage, the LV myocardium was delineated within each frame throughout a single cardiac cycle (end-diastole to end-systole). A black-and-white mask delineated the region of interest for the myocardium within the echocardiographic image for the two apical views. From the segmented myocardium, we subsequently delineated the LV segments as per the conventional AHA-defined myocardial segments [26] for the a2c and a4c views, using an automated computerized algorithm. The original image, together with its corresponding binary myocardial segments, was then fed to a radiomics pipeline for the extraction of 2D shape-based, 1^st^ order, and texture-based features. The complete automated pipeline was developed using open-source Python (version 3.7, Python Software Foundation) and has been previously validated for extracting ultrasomics features [15].

### Morphology and texture-driven handcrafted radiomics

PyRadiomics (version 3.0.1, Python Software Foundation [28]) and SimpleITK (version 2.2.0, Insight Software Consortium [29]) employed within the open-source Python framework (version 3.7.13) facilitated the extraction of ultrasomics features. A total of 98 shape, histogram, and texture-based features were extracted for each of the 12 myocardial segments. To capture temporal and spectral variations within the ultrasomics features, a root mean square and spectral entropy for each feature were calculated across all frames within a single cardiac cycle. Subsequently, all the ultrasomics features from a single segment from the four feature categories (98 from end-diastole, 98 from end-systole, 98 temporal, and 98 spectral) were compiled. Ultrasomics features from all 12 myocardial segments were compiled for each patient (total of 2,352 features per view and 4,704 features per patient) and utilized for feature engineering and machine learning model development.

### Deep learning-based radiomics

In this study, we additionally extracted deep learning-based radiomics features [30–32] from the end-diastolic phase of the echocardiography recordings to assess their efficacy compared to handcrafted radiomics (HCR) features. The deep learning-based radiomics features were extracted using a transfer learning approach from the last classification layer of the ResNet50-based convolutional neural network architecture for each left ventricle myocardium. A total of 1000 deep radiomics features, referred to as deep transfer learning (DTL) features, were extracted from each myocardium and compared for their incremental value over a set of 98 handcrafted radiomics (HCR) features. These HCR features were extracted equivalently using PyRadiomics (version 3.0.1, Python Software Foundation [28]) from the end-diastolic phase for each myocardium within both A2C and A4C views. Using DTL features, we developed an ML model identifying infarcted and non-infarcted LV myocardium. This development aimed to assess any potential improvement in overall performance resulting from the utilization of both types of ultrasomics features. As a preprocessing step, the data from each of the ultrasomics features was normalized using the cumulative distribution function making the data within the scale of 0-1 for all features [15].

### Machine learning model development

Following feature extraction, we developed a patient-level ML-based model (Figure 1B) to differentiate infarcted myocardium from non-infarcted myocardium for each patient (an individual-level model that included data from both views) and a view-level model (Figure 1C) that identified infarcted myocardium when presented with any of the two apical views. Note that the patient-level model provided the infarction label for each patient, whereas the view-level model provided the infarction label for each view within a patient. The machine learning model development was performed using an automated machine learning (AutoML) pipeline provided by the open-source H2O platform (H2O.ai, version 3.44.0.2 [33]).

The AutoML platform utilizes a series of ML algorithms [33], including generalized linear models, deep neural networks, distributed random forests, gradient boosting machines, and extreme gradient boosting. Each of these algorithms was trained on a training dataset (n=296) using a 10-fold cross-validation approach. This approach minimizes the effect of overfitting by assessing the model performance on diverse subsets of the original training data and selecting the optimal model. The external validation cohort (n=84) was then used as a holdout set to evaluate the performance of the optimal model. A similar approach was adopted to develop both the patient-level and view-level ML models. The prediction probabilities from both the patient-level and view-level models were on a 0-1 scale and served as a continuous ultrasomics infarction score. They can also be interpreted as a binary outcome, with any value ≥ 0.5 indicating the presence of infarction, and a value < 0.5 indicating the absence of infarction within a patient or view.

### Performance evaluation and statistical analysis

Baseline clinical characteristics are presented as median (interquartile range) for continuous variables and total count (percentage) for categorical variables. Continuous variables in infarcted and non-infarcted groups are compared using an independent sample t-test if the variable is normally distributed in the two groups or using the Mann-Whitney U test otherwise. Categorical variables across the two groups are compared using Fisher’s exact test if the value in the contingency table is less than 5 or using the chi-square test otherwise. Similarly, ultrasomics from infarcted and non-infarcted segments within each patient for the entire cohort was tested using a paired Mann-Whitney U test. The overall performance of the ML model on the training dataset is evaluated using 10-fold cross-validation and expressed as mean ± standard deviation of five performance evaluation metrics such as sensitivity, specificity, accuracy, F1-score, and area under the curve (AUC), obtained in each of the ten folds. The area under receiver operating characteristics curves was estimated using the DeLong test. Similar metrics were used to demonstrate the generalizability of the model on the external test set. All machine learning evaluations and statistical analyses were performed using Python (version 3.7) and MedCalc (version 12.5.0.0) respectively. A p-value < 0.05 was considered significant.

## Results

### Baseline characteristics

The baseline characteristics of the participants in the training dataset from the HMC-QU database have been previously published [20, 21, 34]. Characteristics of healthy non-infarcted subjects with and without cardiac risk factors are shown in the supplementary material (Table S1). Table 1 shows the baseline characteristics of the participants in the test dataset that included the DTU-STEMI trial [23] and non-infarcted patients. In both the infarcted and non-infarcted groups of patients in the test dataset, age and sex were reported to be significantly different (p<0.001), with patients in the infarcted group being slightly older compared to the non-infarcted group (58 vs 51 years, p<0.001). Considering baseline echocardiographic parameters, patients with myocardial infarction (MI) were reported to have significantly reduced left ventricular ejection fraction (49.9% vs. 62%, p<0.0001), increased relative wall thickness (0.43 vs. 0.36 mm, p<0.0001), higher wall motion score index (p<0.0001), and reduced global longitudinal strain (-12.23% vs. -16.80%, p<0.0001) compared to non-infarcted controls.

**Table 1.** Baseline characteristics for the external test dataset that includes participants from STEMI-DTU trial and MT-Sinai studies. A = late diastolic transmitral flow velocity; BMI = body mass index; E = early diastolic transmitral flow velocity; e’ = early diastolic relaxation velocity at septal mitral annular position, GLS = global longitudinal strain; LVEF = left ventricular ejection fraction; LVEDV = left ventricular end-diastolic volume; LAVi = left atrial volume index; LVMi = left ventricular mass index; WMSI = wall motion score index;

| Variable | Missing | Overall (n=84) | MI (n=40) | Controls (n=44) | p-value |
| --- | --- | --- | --- | --- | --- |
| Age, yrs | 0 (0.0) | 53 (49 - 62) | 58 (52 - 68) | 51 (43 - 56) | <0.001 |
| Sex (Male) | 0 (0.0) | 47 (56) | 31 (78) | 16 (36) | <0.001 |
| BMI, kg/m<sup>2</sup> | 0 (0.0) | 27.26 (23.98 - 33.07) | 27.47 (24.42 - 32.81) | 26.80 (23.27 - 33.42) | 0.34 |
| Smoking | 0 (0.0) | 35 (41.7) | 20 (50.0) | 15 (34.1) | 0.21 |
| Diabetes mellitus | 0 (0.0) | 11 (13.1) | 6 (15.0) | 5 (11.4) | 0.87 |
| Hypertension | 0 (0.0) | 33 (39.3) | 19 (47.5) | 14 (31.8) | 0.21 |
| Heart Rate, bpm | 0 (0.0) | 72.90 (63.30 - 86.50) | 81.00 (73.75 - 93.50) | 66.05 (57.92 - 72.80) | <0.0001 |
| LVEF, % | 2 (2.4) | 58.50 (50.10 - 64.75) | 49.90 (40.75 - 53.08) | 62.00 (59.00 - 66.00) | <0.0001 |
| E, m/s | 0 (0.0) | 0.70 (0.60 - 0.86) | 0.69 (0.62 - 0.90) | 0.73 (0.58 - 0.83) | 0.54 |
| A, m/s | 0 (0.0) | 0.66 (0.51 - 0.79) | 0.73 (0.56 - 0.82) | 0.60 (0.47 - 0.74) | **0.032** |
| E/A | 0 (0.0) | 1.10 (0.88 - 1.40) | 1.09 (0.87 - 1.21) | 1.20 (0.90 - 1.40) | 0.15 |
| Average e', cm/s | 13 (15.5) | 8.00 (6.33 - 10.29) | 6.60 (5.49 - 7.68) | 9.76 (7.37 - 11.25) | <0.0001 |
| E/e' | 14 (16.7) | 8.92 (6.99 - 12.16) | 12.26 (7.89 - 15.88) | 7.96 (6.12 - 10.07) | <0.001 |
| LVEDV, mL | 5 (6.0) | 97.08 (84.24 - 113.50) | 94.89 (87.11 - 126.68) | 98.05 (81.70 - 112.25) | 0.77 |
| LVESV, mL | 5 (6.0) | 35.86 (29.55 - 47.88) | 42.95 (33.60 - 55.83) | 33.35 (28.38 - 38.12) | **0.001** |
| LAVi, mL/m<sup>2</sup> | 17 (20.2) | 29.70 (24.75 - 34.15) | 28.31 (22.51 - 33.94) | 30.05 (26.05 - 34.70) | 0.22 |
| LVMi, g/m<sup>2</sup> | 17 (20.2) | 72.53 (61.25 - 83.69) | 85.93 (73.82 - 98.40) | 64.97 (59.23 - 75.09) | <0.0001 |
| WMSI | 18 (21.4) | 1.00 (1.00 - 1.00) | 1.83 (1.06 - 2.06) | 1.00 (1.00 - 1.00) | <0.0001 |
| GLS, % | 24 (28.6) | -15.01 (-17.33 - -11.10) | -12.23 (-15.13 - -9.67) | -16.80 (-18.96 - -14.89) | <0.0001 |

### Prediction of myocardial infarction

In this study, we developed the patient-level and view-level ML models using the autoML pipeline for identifying patients with and without MI. The rigorous training of the AutoML pipeline resulted in the selection of the gradient boost model (XGBoost) as the optimal choice for MI prediction, with a set of hyperparameters provided in the supplemental material (Table S2). This model was then evaluated for performance on the external test dataset. The patient-level ML model for predicting MI showed an average 10-fold cross-validation sensitivity of 88.15±8%, specificity of 85.23±13.7%, F1-score of 81.85±9.5%, accuracy of 86.07±8.9%, and AUC of 0.88±0.07. In the external test dataset, the model showed a sensitivity of 78.95%, a specificity of 84.78%, an F1-score of 80%, an accuracy of 82.17%, and an AUC of 0.93 (95% CI: 0.87-0.97) (Figure 2).

**Figure 2.**
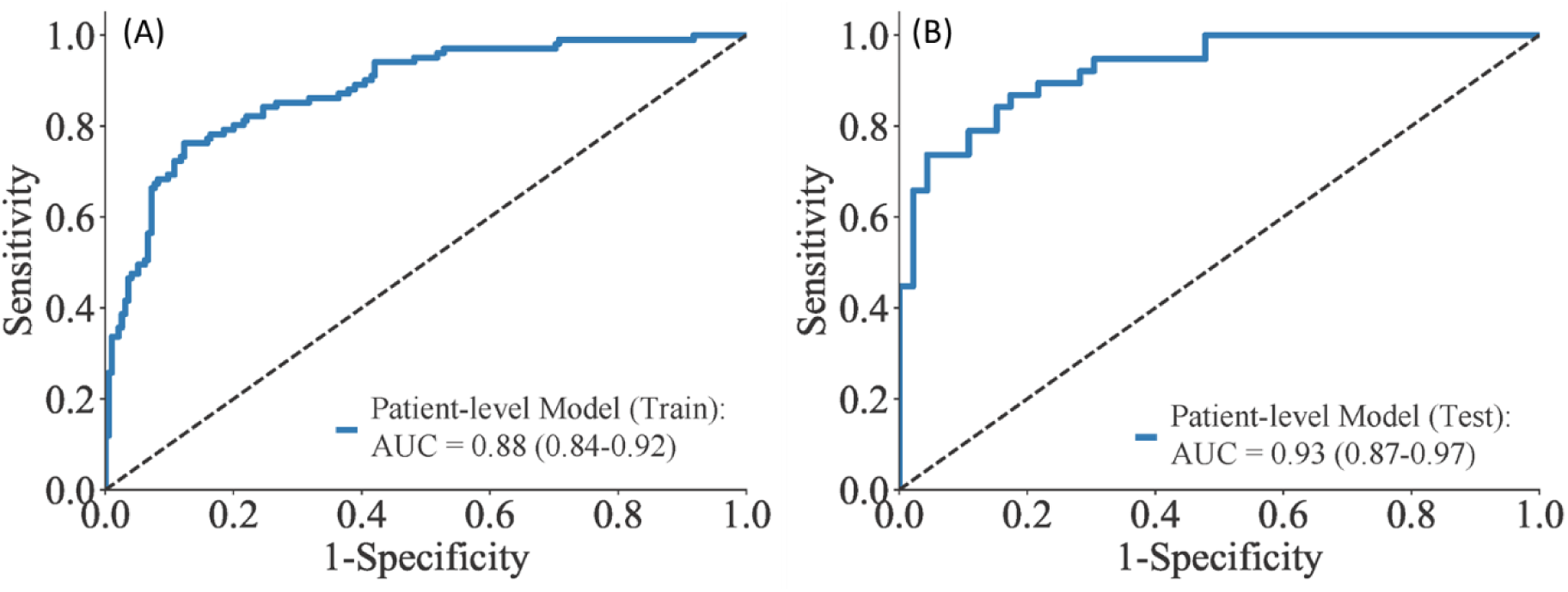
Performance of the patient-level ML model for predicting the MI in (A) training dataset and (B) external test dataset.

For the view-level model, the training cross-validation showed an average sensitivity of 84.15±9.4%, specificity of 86.53±9.8%, F1-score of 79.24±6.2%, accuracy of 85.54±5%, and AUC of 0.90±0.04. Whereas on the external test dataset, the model demonstrated a sensitivity of 49%, specificity of 92%, F1-score of 61%, accuracy of 75%, and an AUC of 0.83 (95% CI: 074-0.89) (Figure 3).

**Figure 3.**
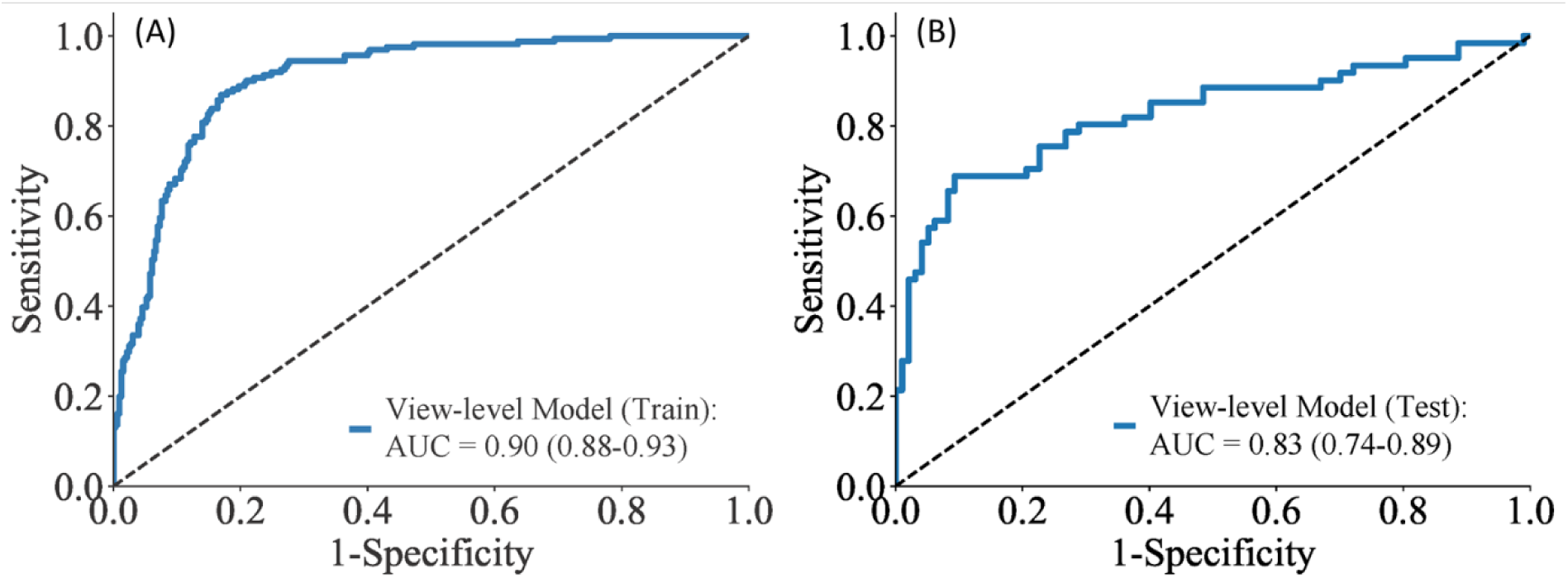
Performance of the view-level ML model for predicting the MI in (A) the training dataset and (B) the test dataset.

### Comparison of handcrafted radiomics with deep radiomics for predicting MI

We investigated the efficacies of HCR and DTL features independently and jointly using machine learning models (Figure 4). The 10-fold cross-validation performance on the training set reported an average AUC of 0.84±0.04 for the HCR model, 0.81±0.06 for the DTL model, and 0.83±0.07 for the combined model with both HCR and DTL features. Our findings revealed that on the external test dataset, the ML model developed using DTL features demonstrated equivalent performance (AUC = 0.75, 95% CI: 0.66-0.82) in identifying infarcted myocardium compared to the ML model developed using HCR (AUC = 0.74, 95% CI: 0.66-0.82). However, when both HCR and DTL features were combined, a slight improvement in performance in terms of AUC was observed (AUC = 0.79, 95% CI: 0.71-0.85).

**Figure 4.**
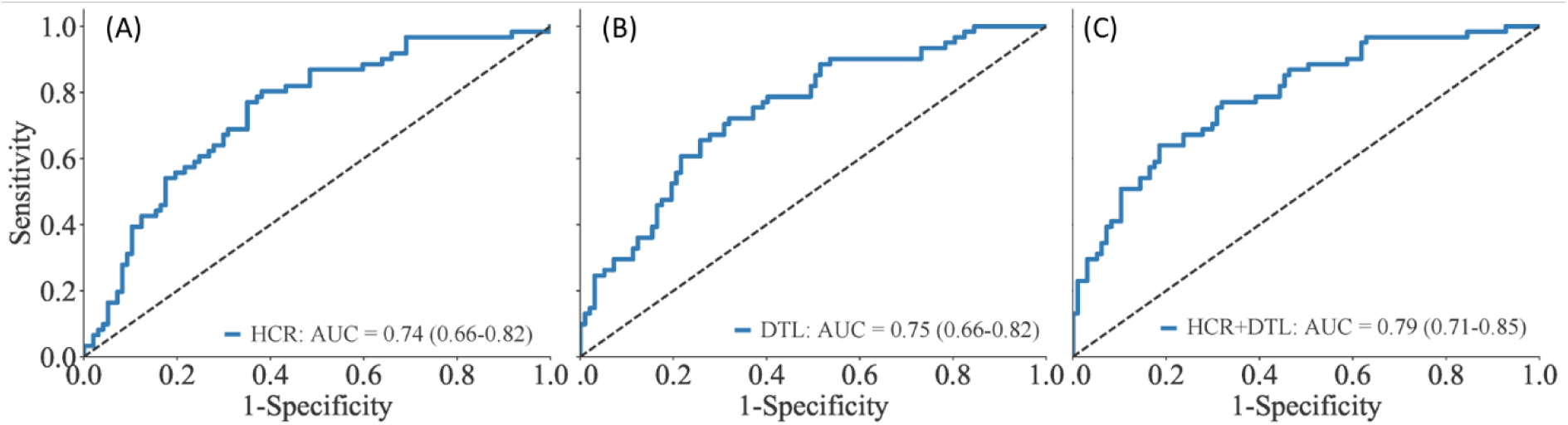
Comparing the performance of ML models developed using (A) handcrafted radiomics (HCR), (B) deep transfer learning-based radiomics (DTL), and (C) a combination of HCR and DTL in identifying the infarcted myocardium. HCR = handcrafted radiomics; DTL = transfer learning-derived deep learning-based radiomics.

### Parametric visualization of infarcted myocardium

For parametric radiomics visualization over the myocardium in each view, we used the HCR model as discussed in the previous section (Figure 4 A) and obtained the SHapley Additive exPlanations (SHAP) plot from the model (Figure 5). The SHAP plot provided the overall contribution of ultrasomics features in identifying the infarcted myocardium. We identified a texture feature (gray-level nonuniformity feature, extracted from the gray-level dependence matrix) as highlighted in the SHAP plot (Figure 5). This feature also demonstrated a significant difference (p=0.0007) when pairwise comparisons were performed between infarcted and non-infarcted segments within each patient (Table S3). We generated a parametric feature map to visualize variations in gray level intensities within the LV myocardium on each echocardiographic view. Figure 6 displays the close correspondence of the delayed gadolinium-enhanced CMR and Echo images for localizing the regions of infarcted myocardium over the anterior wall in the patients from the DTU-STEMI. For example, as shown in Figure 6 A and Figure 6 B, the infarct is situated within the apical inferior, apical anterior, and mid-anterior segments of an MI patient, as evident in both the CMR scan and a corresponding 2-chamber echocardiographic view. Conversely, in healthy subjects without infarcted segments, none of the segments were highlighted (Figure 6 C).

**Figure 5.**
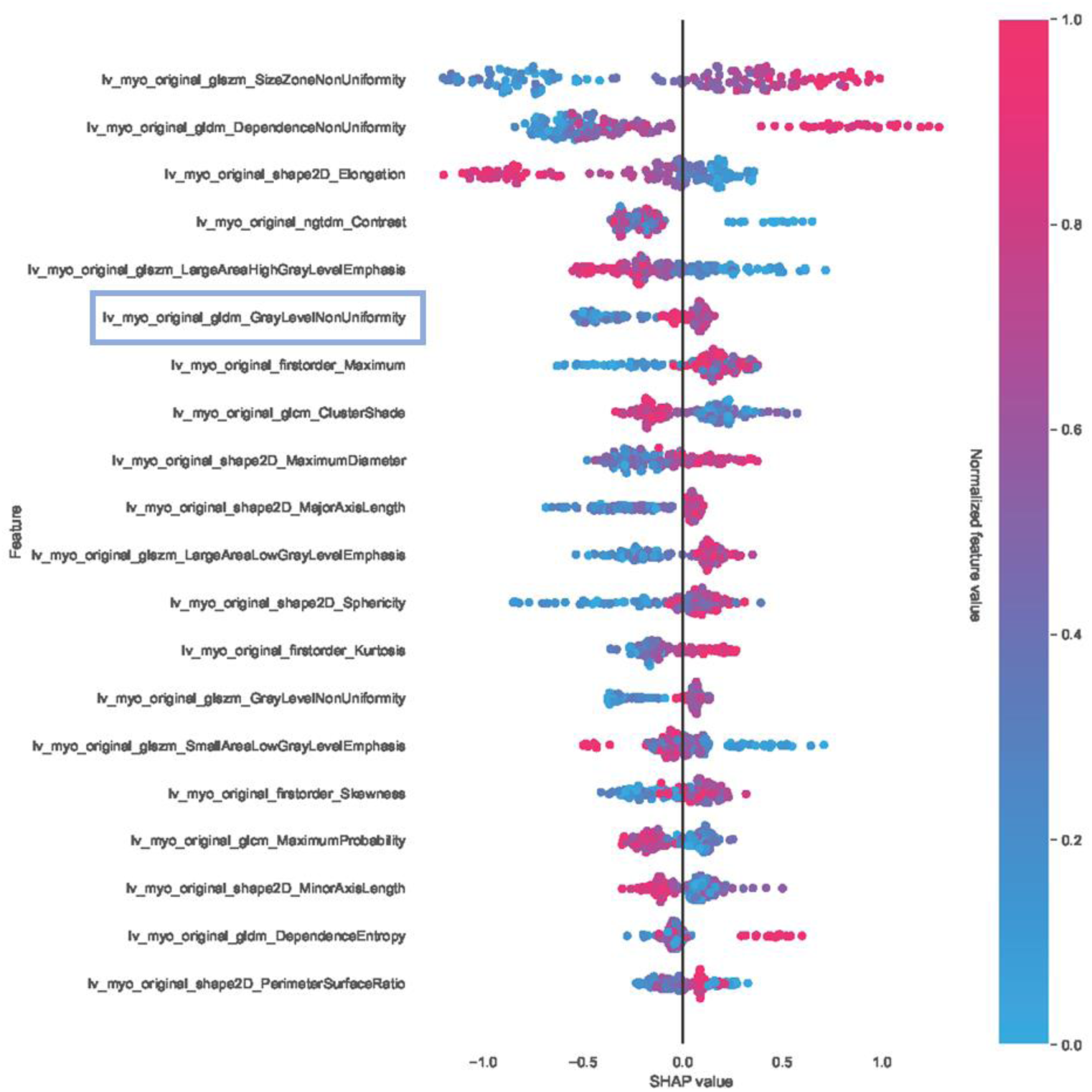
SHAP plot for the MI prediction model.

**Figure 6.**
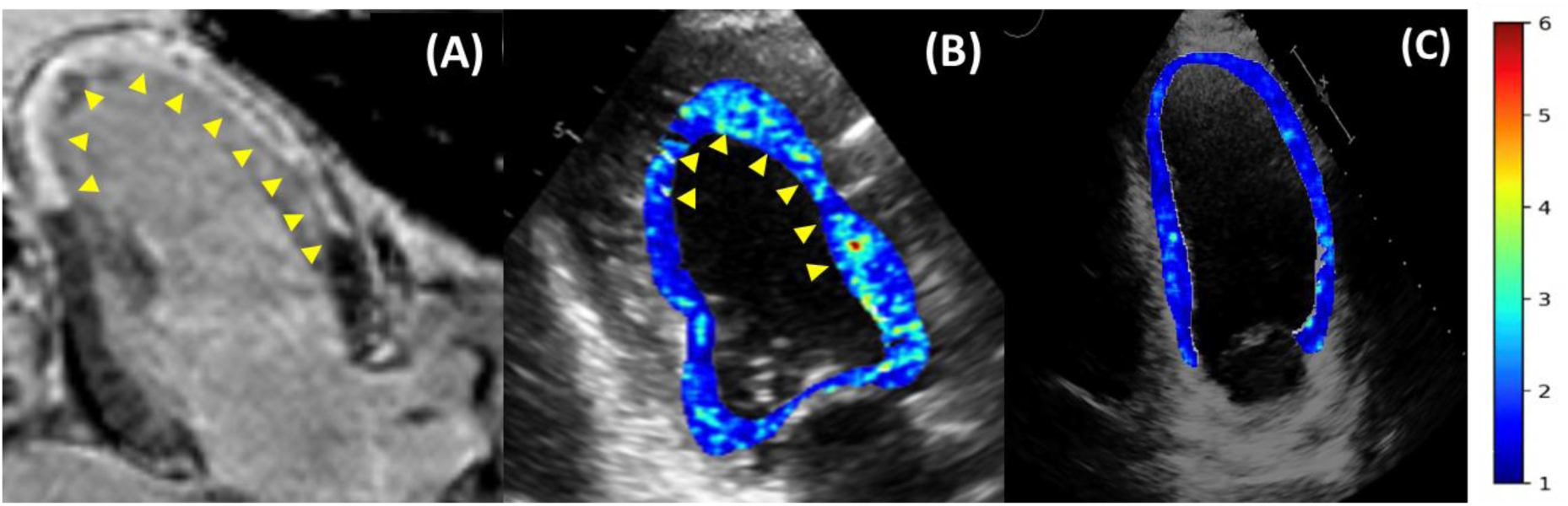
Visualization of ultrasonic texture feature maps over the LV myocardium: Panels A-B show CMR and corresponding 2-chamber echocardiographic views for patients with myocardial infarction. Panel C illustrates a participant without infarcted segments.

## Discussion

CMR is currently the gold standard for infarct size quantification in clinical trials [6, 35]. However, the widespread use of CMR is somewhat limited by cost and availability. Therefore, exploring the role of radiomics for the direct identification of infarcted tissue may aid the future development of cost-effective strategies to estimate infarct size using cardiac ultrasound techniques. To this end, this study explored the utility of characterizing ultrasonic texture changes using radiomics for predicting the presence of infarcted myocardium. Machine learning models using cardiac ultrasound radiomics, also referred to as ultrasomics, showed good discrimination in identifying patients with MI. Moreover, the ability of radiomics to delineate infarcted myocardium using individual end-diastolic frames and the potential to delineate infarcted myocardium using parametric display illustrates the promise of radiomics approaches for facilitating cardiac ultrasound imaging-based myocardial tissue characterization.

Individual structural components of the myocardium influence its acoustic properties [36]. Infarcted myocardium is characterized by stretching and rearrangement of necrotic myocytes, surrounded by contracted myocytes, edema, hemorrhage, and inflammatory reparative processes [37]. These changes within the myocardium influence ultrasound signal intensity distributions [38, 39]. The present work extends our previous observations [16] where specific analyzable trends in ultrasound texture information were seen to identify tissue-based changes in cardiac remodeling including the prediction of patients who showed the presence of LGE in myopathic hearts. However, in contrast to previous studies we show the feasibility of radiomics features to be extracted even from apical echocardiography views for developing machine learning models. Second, we extracted radiomics features over the entire cardiac cycle and showed that the incorporation of cyclical variance in radiomics features improved the model discrimination. This cyclical variance of radiomics features may represent an attribute of regional function. Further work is needed to directly test its relationship with wall motion analysis and incremental value over regional and global LV strain.

Convolutional Neural Networks (CNNs) stand out as the most popular deep learning algorithms utilized extensively in medical image analysis [17, 40–42]. Their widespread adoption is primarily because of their inherent ability to autonomously extract patterns or features from images without relying on external handcrafted features, a requirement common in conventional machine learning algorithms. Many CNNs have been trained on large ImageNet datasets, showcasing promising results when applied to medical image tasks through the transfer learning approach [43]. In our study, we also employed this transfer learning approach to extract deep learning-based features using the ResNet50 architecture. By combining radiomics with deep features, we tried to enhance generalizability by merging high-level representations and domain-specific knowledge. However, our results show that deep radiomics alone may not surpass handcrafted radiomics in predicting myocardial infarction. This could be due to the ability of handcrafted radiomics to capture intricate tissue-level changes more effectively. Our results also showed no significant improvement in predictive performance with this combined approach. This lack of improvement may be attributed to the limited sample size utilized for training the CNN models. It is noteworthy that CNNs generally demonstrate better performance with larger sample sizes [44]. Additionally, our feature extraction process focused solely on end-diastolic frames, potentially constraining the model’s ability to capture the full spectrum of relevant features. These results suggest that handcrafted radiomics may offer equivalent performance under these circumstances. However, to achieve superior performance, increasing the sample size and incorporating a wider range of features are crucial.

Several limitations of the present study and opportunities for future research need to be acknowledged. First, including both STEMI and non-STEMI patients may allow testing the model performance over a range of infarct sizes. Second, the apical 3-chamber view was not available in the publicly available training dataset and the inclusion of apical 3-chamber views would be necessary for comprehensive evaluation. Third, we did not directly test the ability of radiomics to predict the actual CMR-derived infarct size. This would require the development of regression algorithms that not only display the infarcted region but also estimate the overall size and infarct location. The infarcted region is heterogeneous and includes a range of tissue characteristics ranging from necrotic, ischemic yet viable tissue, and microvascular obstruction to intramyocardial hemorrhage. The differential effects of these features impact the future recovery of myocardial function and would need more in-depth analysis. Finally, the prognostic importance of infarct size delineated using cardiac ultrasound radiomics would require to be tested. The use of more complex designs and exploring these questions in future prospectively designed studies using direct head-to-head comparison with CMR could yield more insights into the future potentials of this technique.

## Conclusion

In summary, our study explored patient-level and view-agnostic machine learning models that leverage ultrasomics features for identifying patients with MI and infarcted myocardium. This preliminary data underscores the potential of pixel-level statistical and texture patterns for myocardial tissue characterization using cardiac ultrasound images.

## Data Availability

All data produced in the present study are available upon reasonable request to the authors.

