## Supplemental material for "Cardiac Ultrasonic Tissue Characterization in Myocardial Infarction Based on Deep Transfer Learning and Radiomics Features"

**Supplementary Material**

Table S1. Clinical characteristics for the WVU database.

| Variable | Overall (n=134) | Missing |
|---|---|---|
| **Age, yrs** | 37.00 (27.00 - 51.00) | 1 (0.7) |
| **Sex (Male)** | 64 (47.8) | 0 (0.0) |
| **Body mass index** | 26.28 (23.34 - 31.74) | 0 (0.0) |
| **Diabetes mellitus** | 9 (6.7) | 0 (0.0) |
| **Hypertension** | 34 (25.4) | 0 (0.0) |
| **Heart rate** | 73.00 (61.00 - 81.25) | 6 (4.5) |
| **Hyperlipidemia** | 39 (29.1) | 0 (0.0) |
| **CAD** | 0 (0.0) | 0 (0.0) |
| **PAD** | 1 (0.7) | 0 (0.0) |
| **Obesity** | 40 (29.9) | 0 (0.0) |
| **Valvular Heart Disease** | 0 (0.0) | 0 (0.0) |
| **CKD** | 0 (0.0) | 0 (0.0) |
| **Asthma** | 16 (11.9) | 0 (0.0) |
| **COPD** | 4 (3.0) | 0 (0.0) |
| **DVT or PE** | 3 (2.2) | 0 (0.0) |
| **Anemia** | 3 (2.2) | 0 (0.0) |
| **Cancer active** | 6 (4.5) | 0 (0.0) |
| **Cancer remission** | 8 (6.0) | 0 (0.0) |
| **ETOH Abuse** | 14 (10.4) | 0 (0.0) |
| **Illicit Drug Use** | 3 (2.2) | 0 (0.0) |
| **LVEDV, mL** | 94.95 (81.47 - 113.25) | 2 (1.5) |
| **LVESV, mL** | 35.60 (27.50 - 43.00) | 7 (5.2) |
| **LVEF, %** | 63.00 (59.85 - 66.75) | 0 (0.0) |
| **LVMi, g/m2** | 58.92 (51.13 - 72.02) | 0 (0.0) |
| **E, m/s** | 0.80 (0.71 - 0.91) | 3 (2.2) |
| **A, m/s** | 0.58 (0.46 - 0.74) | 3 (2.2) |
| **E/A** | 1.42 (1.09 - 1.81) | 3 (2.2) |
| **Average e', cm/s** | 11.80 (9.78 - 14.00) | 4 (3.0) |
| **Average E/e'** | 7.16 (5.86 - 8.52) | 7 (5.2) |
| **TRVmax** | 1.95 (1.60 - 2.21) | 63 (47.0) |

CAD = coronary artery disease; PAD = peripheral artery disease; CKD = chronic kidney disease; COPD = Chronic obstructive pulmonary disease; DVT/PE = deep vein thrombosis & pulmonary embolism; ETOH = ethyl alcohol or ethanol; LVEDV = left ventricular end-diastolic volume; LVESV = left ventricular end-systolic volume; LVEF = left ventricular ejection fraction; LVMi = left ventricular mass index; A = late diastolic transmitral flow velocity; BMI = body mass index; E = early diastolic transmitral flow velocity; e' = early diastolic relaxation velocity at septal mitral annular position, TRV = tricuspid regurgitation velocity

Table S2 The hyperparameters of the patient-level XGBoost model.

| Parameter | Value |
|---|---|
| **Number of trees** | 50 |
| **Maximum depth** | 6 |
| **Minimum child weight** | 1 |
| **Learning rate** | 0.6 |
| **eta** | 0.3 |
| **Sample rate** | 1 |
| **Subsample** | 1 |
| **Maximum bins** | 256 |

Table S3 A pairwise comparison to identify the features that discriminate the infarcted segments as against the non-infarcted segments within a view.

| Ultrasomic Features | p-value |
|---|---|
| NGTDM Strength at ED frame | p<0.0001 |
| NGTDM Coarseness at ED frame | p<0.0001 |
| GLSZM Large Area High Gray Level Emphasis at ES frame | 0.00012 |
| NGTDM Strength at ES frame | 0.0002 |
| NGTDM Coarseness from spectral variations | 0.0005 |
| GLDM Gray Level Non-Uniformity from temporal variations | 0.0007 |
| NGTDM Coarseness from temporal variations | 0.0007 |
| GLDM Gray Level Non-Uniformity from ED frame | 0.0007 |

NGTDM: Neighbouring Gray Tone Difference Matrix, GLSZM: Gray Level Size Zone Matrix, GLDM: Gray Level Dependence Matrix
